# Bridging Genomics and Pharmacoepidemiology to Expand Treatment Options for Alcohol Use Disorder

**DOI:** 10.64898/2026.01.09.26343615

**Authors:** Christopher T. Rentsch, Samantha G. Malone, Mingjian Shi, Michael R. Setzer, Zachary Piserchia, Emma L. Winterlind, Mehdi Farokhnia, John Tazare, Amy C. Justice, David A. Fiellin, Lorenzo Leggio, Henry R. Kranzler, Joshua C. Gray

**Author notes:** **Corresponding author:** Joshua C. Gray, PhD.

## Abstract

Alcohol use disorder (AUD) is a chronic, relapsing condition and a major public health problem. However, few medications are approved to treat AUD, and those available show limited efficacy. Drug repurposing is a cost-effective strategy to identify novel therapeutic uses for existing medications. Here, we describe a pipeline that integrates genetic and electronic health record (EHR) data to identify and evaluate drugs to be repurposed for treating AUD. Our approach comprises 1) alcohol-associated gene identification and biological network generation; 2) mapping drugs to target proteins; 3) filtering promising repurposing candidates; and 4) an exemplar pharmacoepidemiologic analysis of the effect of an identified drug (i.e., baclofen) on alcohol consumption. Linking loci to genes from a genome-wide association study (GWAS) of problematic alcohol use identified 94 genes, which we expanded to 327 alcohol-related genes through network-based analyses. Across these analyses, 52 genes were linked to 195 FDA-approved drugs, including four already approved or used off-label to treat AUD. After filtering for safety, relevance, and data availability, 26 candidate drugs, including baclofen, were selected for further evaluation. An evaluation of the real-world effectiveness of baclofen using national EHR data from the United States Department of Veterans Affairs provided evidence that baclofen-exposed patients reduced alcohol consumption more than propensity-score-matched unexposed patients. This approach, which aligns genomic findings with real-world clinical data, provides an efficient method for identifying promising drug repurposing candidates and prioritizing those that merit evaluation in randomized trials to ultimately advance pharmacotherapies for AUD.

## Introduction

Alcohol use disorder (AUD) is a chronic, relapsing condition characterized by a preoccupation with alcohol and an impaired ability to stop or control alcohol use despite adverse social, occupational, or health consequences^1^. In 2023, approximately 28.1 million adults in the U.S. experienced AUD in the past year^2,3^. Despite the debilitating impact of AUD on public health, only three medications (i.e., disulfiram, naltrexone, and acamprosate) are approved by the Food and Drug Administration (FDA) for treating AUD. Of concern, only a minority of patients with AUD receive these treatments ^4^ and among those who do, there is heterogeneity in patient response^5–9^. Prescriber knowledge of and comfort with drugs to treat AUD is a known barrier to their use^10,11^.

Developing new medications for complex neuropsychiatric disorders like AUD is challenging. Only 15% of medications that enter clinical trials for brain diseases are approved by the FDA for marketing^12^. Across therapeutic areas, estimated medication development costs increase nearly 9% above inflation annually^13^, with current costs reaching $1-3 billion per drug^14,15^. Repurposing drugs approved for other indications and with known safety profiles, could dramatically reduce research and development costs^16^. For example, prior rigorous testing of approved medications in a wide range of patients during Phase III trials and ongoing surveillance in Phase IV trials greatly lower the chances of unanticipated adverse events with repurposed drugs. Medications prescribed by clinicians for other conditions would also have the benefit of previously established prescriber knowledge and comfort.

Historically, drug repurposing has involved clinical observation and chance discoveries. However, recent efforts have used systematic computational approaches to leverage genomic biobank and electronic health record (EHR) data to identify candidates for repurposing^15,17^.

Leveraging existing genomic data holds value because targeting disease mechanisms with genetic support increases the success rate of medication development over two-fold^18,19^. Thus, genes that affect alcohol consumption or risk of problematic alcohol use (PAU; i.e., AUD diagnosis or symptomatology) could yield new targets and medications for therapeutic repurposing. Biological networks can be used to extend genetic discoveries by identifying additional proteins that interact with disease-associated genes. These networks are often enriched for targets of approved medications^20,21^, suggesting that genes linked to alcohol consumption and PAU—as well as their broader biological context—may implicate novel pharmacotherapeutic targets.

The increasing availability of large EHR databases and routine collection of alcohol consumption data enable retrospective analysis of the real-world effects of promising medications on alcohol consumption using advanced pharmacoepidemiologic methods that can partially address the biases inherent in observational studies^22^. Such analyses assess promising drugs and aim to prioritize top candidates for evaluation in preclinical studies and randomized clinical trials (RCTs)^15^. Studies that use real-world data can yield conclusions comparable to those from RCTs when appropriate designs and analyses are employed^23^. Indeed, our team has examined several medications for treating AUD, using designs that balance treatment groups with statistical methods analogous to randomization in trials. To date, we have found support for gabapentin, spironolactone, topiramate, and glucagon-like peptide-1 receptor agonists (GLP-1RAs)^24–29^, with findings from these studies informing and guiding ongoing clinical trials (NCT06015893 and NCT05807139).

Building on this work, we have developed a comprehensive pipeline integrating genomic-driven discovery with real-world data analysis within the Department of Veterans Affairs (VA), the largest integrated healthcare system in the United States (∼9 million patients served annually)^30^. The aim of this paper is to develop and describe a data pipeline that integrates genetic and EHR data to identify potentially novel treatments for AUD. We hypothesized that this pipeline would identify multiple drugs whose targets are genetically implicated in AUD and therefore represent promising candidates for repurposing. Below we outline the methodology (Figure 1) and results for 1) alcohol-associated gene identification and biological network generation; 2) drug-protein identification; 3) filtering promising drugs for repurposing; 4) an exemplar pharmacoepidemiologic analysis of the effect of an identified drug candidate (i.e., baclofen) on alcohol consumption in the VA EHR.

## Methods

The methods are organized to mirror the sequential steps of the analytic pipeline introduced above and depicted in Figure 1. Briefly, we (1) identified genetically informed targets for AUD; (2) mapped FDA-approved drugs to these targets; (3) filtered candidate drugs based on clinical suitability and feasibility for pharmacoepidemiologic analysis; and (4) conducted an exemplar pharmacoepidemiologic analysis to evaluate one prioritized candidate (baclofen) using VA EHR data. Each subsection below corresponds to one step of this pipeline.

## Selecting genetically informed targets

The initial steps of our pipeline were to utilize GWAS and biological network generation to identify relevant gene targets related to alcohol use. Loci from a GWAS of PAU^31^ were assigned to genes based on either their presence within the gene (determined using dbSNP [https://www.ncbi.nlm.nih.gov/snp/]) or proximity to the nearest gene transcription start site (obtained from Open Targets [https://genetics.opentargets.org/ v22.10;^32^ ]). Of note, a recent benchmarking study found that a simple nearest gene method performed the same or better than more complex methods such as expression quantitative trait locus colocalization^33^.

Second, alcohol-related networks were derived from the output of a recently developed and validated method for the network-based expansion of causal genes^20^. The algorithm took as input likely causal seed genes (>0.5 posterior probability from the locus-to-gene algorithm from Open Targets Genetics)^34^ at each genome-wide significant locus from 1,002 EFO terms, including alcohol-related GWAS focused on in this study (EFO_0007878: alcohol consumption measurement; EFO_0004329: alcohol drinking; EFO_0003829: alcohol dependence; EFO_0007835: alcohol dependence measurement). The genes for each trait were run separately, propagating through an interactome (see ^20^ for more detail) using the Personalized PageRank (PPR) algorithm from the R package igraph (v.1.2.4.2). The algorithm selected nodes with a PPR ranking score above the third quartile (Q3, 75%) and applied the walktrap clustering algorithm (igraph v.1.2.4.2) to generate modules. If a module contained more than 300 nodes, additional clustering was performed within that community until all resulting clusters contained fewer than 300 genes. The top alcohol-related network was prioritized based on the Kolmogorov-Smirnov test *p*-value (i.e., an indicator of network coherence), the inclusion of FDA-approved drugs for AUD that interact with the network genes, and the theoretical relevance of seed and peripheral genes. Theoretical relevance was assessed based on biological plausibility—specifically, whether genes mapped onto pathways, receptors, or mechanisms previously implicated in alcohol use or AUD, including those targeted by FDA-approved or guideline-recommended medications, or other proteins with well-documented roles in alcohol-related neurobiology. The full description of the interactome, algorithmic implementation, code, and network-generation results—including the full set of alcohol-related networks—may be found in the original repository (https://zenodo.org/records/7575743) and publication ^20^. We did not modify or rerun the algorithm; rather, we relied on the alcohol-related networks reported in the supplemental tables of the original manuscript.

## Drug-protein identification

Medications were linked to genes utilizing the Open Targets Platform “Target” dataset (https://platform.opentargets.org/downloads). For each gene, the ‘Known Drugs’ field aggregates drug–target relationships curated through ChEMBL^35^ .The ChEMBL database curates compounds, activity measurements, assays, and targets from the scientific literature using both manual and semi-automated methodologies. The curation process enhances data accessibility, ensures comparability through standardization, and improves reliability by identifying outliers, obscurities, and errors. It also adds annotations and mappings to increase data accuracy (for more information see^36,37^). We restricted analyses to drugs with documented, curated interactions with human proteins and filtered to include only FDA-approved medications.

## Filtering of drugs for clinical suitability

Drug filtering was based on two criteria: 1) clinical suitability and 2) feasibility for pharmacoepidemiologic analyses. Filtering was conducted by ELW and JCG in consultation with two physicians (HRK, LL) who oversaw the filtering process, informed the exclusion criteria, and approved the final list of drugs and consolidation of drugs into medication groups for testing based on identical or near-identical mechanisms of action and treatment indications.

Drug indications and side effects were identified using a combination of Open Targets annotations and FDALabel, from which approved indications and clinically relevant adverse effects were manually extracted^38^. Automated resources such as ChEMBL^35^ (for indications) and SIDER^39^(for side effects) were considered but not used as primary sources due to heterogeneity across contributing registries. Drugs were filtered to eliminate those with severe adverse effects or addiction risk, those used principally as-needed or short-term, and those used exclusively to treat severe impairing diseases (Alzheimer’s, Parkinson’s, multiple sclerosis), rare diseases, infant/pediatric/natal conditions, cancer, chronic kidney disease, myocardial infarction, overdose, narcolepsy, and insomnia.

## Assessing feasibility for pharmacoepidemiologic analyses

To determine feasibility for pharmacoepidemiologic analyses in the VA EHR, we identified the number of unique patients with at least one dispensed prescription of our previously identified drugs between Jan 1, 2008 and Dec 31, 2023 and reported any alcohol consumption at the time of medication receipt. Alcohol consumption was measured by a score > 0 on the Alcohol Use Disorders Identification Test - Consumption (AUDIT-C), a self-reported 3-item questionnaire on alcohol use frequency and quantity that is widely used and validated as a screening tool^40,41^. We consolidated drugs into groups for potential pharmacoepidemiologic analyses based on identical or near-identical mechanisms of action and treatment indications.

Finally, we excluded drug groups initiated by fewer than 1,820 patients to ensure sufficient power for subgroup analyses comprising as little as 10% of the final sample. This threshold was based on a sample size calculation assuming α=0.05, 80% power, an effect size of 0.5, and a standard deviation of 3.2 (based on prior work^25^).

## Exemplar Pharmacoepidemiologic Analysis

From the final set of identified medications, we selected baclofen, a GABA-B receptor agonist that has been used for decades to treat muscle spasticity and has also been investigated as a treatment for AUD. We chose to focus on baclofen because of the inconsistent findings of prior studies in AUD resulting in different recommendations concerning its efficacy and different regulatory decisions across different countries^42–44^. A recent meta-analysis of baclofen’s efficacy in 17 RCTs totaling 1,818 participants compared baclofen (at a daily dosage that varied from 30-300 mg) to placebo or another medication^45^. This meta-analysis showed that baclofen modestly decreased the risk of relapse and increased the percentage of days abstinent, however findings for both outcomes showed substantial heterogeneity. Our analysis aimed to augment prior literature on baclofen as a potential treatment for AUD.

Full methodological details are described in Appendix 1. Briefly, an observational cohort study was conducted, using data from the VA EHR, to examine the association between baclofen receipt (at least 60 continuous days) and change in AUDIT-C scores^41,46^. Each baclofen-exposed patient was propensity-score matched^47,48^ to one unexposed patient, using a greedy matching algorithm^49^. Multivariable difference-in-difference (DiD) linear regression models^50,51^ estimated the differential change between baseline (pre-index) and follow-up (post-index) AUDIT-C scores among exposed and unexposed patients. Subgroup analyses stratified by a diagnosis of current AUD and baseline AUDIT-C score (1-3 = low-risk drinking, 4-7 = at-risk drinking, and ≥ 8 = hazardous/binge drinking).

## Results

### Genetically informed targets

The PAU GWAS identified 100 significant variants at 90 loci in the cross-ancestry meta-analysis. All 100 variants were assigned to genes based on their presence within the gene or proximity to the nearest gene transcription start site, which yielded 94 unique genes. There were 74 alcohol seed genes entered into the network-based expansion algorithm, which generated 20 alcohol-related networks. The top network was prioritized based on the Kolmogorov-Smirnov test p-value, presence of an FDA-approved drug for AUD that interacts with the network genes, and theoretical relevance of seed and peripheral genes. The top prioritized network contained 237 genes, including 6 seed genes, and had the most statistically significant Kolmogorov-Smirnov value of the alcohol-related networks. This network contained seed genes previously implicated in the etiology of AUD (e.g., *DRD2*) and genes linked to approved or unapproved medications with evidence of efficacy in treating AUD (i.e., naltrexone, gabapentin, baclofen). Of the 94 and 237 genes from the respective data sources, 327 (98.8%) were unique (Table S1).

## Drug-protein identification

Five of the 94 genes from the PAU GWAS were targeted by at least one FDA-approved drug. Fifty of the 237 top alcohol-related network genes were targeted by at least one FDA-approved drug. Across both data sources there were 52 unique genes targeted by at least one FDA-approved drug (Table S2). The 52 unique genes were targeted by 195 drugs (Figure 2; Table S3). The PAU gene list implicated topiramate (*GABRA4*) and acamprosate (*GABRA4*), the network gene list implicated naltrexone (*OPRK1* and *OPRD1),* and both gene lists implicated gabapentin (PAU and network: *CACNA1C*; network only: *CACNA1D*, *CACNB2*, *CACNB3*), all medications with evidence of efficacy in treating AUD^52^. Thus, all medications, with the exception of disulfiram, that are recommended in the VA/DOD Clinical Practice Guidelines for treating AUD were identified in this search, supporting the enrichment of this gene list for potential treatment targets^53^.

## Drug filtering for clinical suitability and feasibility

After excluding medications with severe adverse effects, short-term or as-needed use, those approved for treating severe impairing diseases, or those that did not meet the threshold for statistical power, 26 drugs remained for assessment in the VA EHR (Table 1). Filtering the drugs that target genes from the PAU GWAS led to 2 drugs (roflumilast, apremilast) that target the receptor encoded by *PDE4B*. Filtering drugs from the network analysis yielded 17 drugs that target receptors encoded by alpha-adrenergic receptor genes (*ADRA2A*, *ADRA2C*, *ADRA2B*), serotonergic receptor genes (*HTR1A*, *HTR1B*, *HTR1D*), gamma-aminobutyric acid-B receptor genes (*GABBR2*, *GABBR1*), and the dipeptidyl peptidase 4 gene (*DPP4*). Finally, 7 drugs targeted genes that encode calcium channels, including *CACNA1C,* which was implicated by both the PAU and the alcohol-related networks. Consolidating medications with similar mechanisms of action yielded 14 medication groups (Table 1).

## Exemplar pharmacoepidemiologic analysis

Baclofen is presented here as an exemplar candidate to demonstrate the application of the final stage of the analytic pipeline rather than as the sole focus of the study. After applying exclusions for the formal pharmacoepidemiologic analysis (Figure S1), we identified 67,906 baclofen-exposed and 3,049,284 unexposed patients eligible for propensity-score matching. Most (n=66,845; 98.4%) exposed patients were matched to an eligible unexposed patient. Of these, 44,550 (67%) matched exposed and 48,665 (73%) unexposed patients did not have a post-index AUDIT-C score measure and could not be included in the analysis. Thus, the final analytical cohort consisted of 22,295 exposed and 18,180 unexposed patients.

Before propensity score matching, the distribution of baseline characteristics differed between exposed and unexposed patients (Table S4). Compared to the unexposed group, patients who received baclofen were younger (2.2% vs. 15.8% aged ≥80 years), had a higher prevalence of mood disorders (41.0% vs. 15.5%), AUD (19.0% vs. 9.1%), hemiplegia or paraplegia (5.9% vs. 0.3%), and hyperpolypharmacy (10.6% vs. 1.7% with ≥10 chronic medications). Propensity-score matching produced treatment groups that were well balanced (all standardized mean differences ≤0.1). Median follow-up time was 230 days (IQR 135–447) among exposed and 263 days (IQR 135-491) among unexposed patients.

Overall, AUDIT-C scores decreased during the study period in both groups. Average AUDIT-C scores decreased from 2.97 (standard error [SE] 0.02) to 2.09 (SE 0.02) among baclofen-exposed patients and from 2.98 (SE 0.02) to 2.29 (SE 0.02) among unexposed patients (Table 2). Therefore, on average, AUDIT-C scores decreased 0.19 points more among baclofen-exposed patients than unexposed patients (95% CI 0.12 to 0.26; p<0.0001). This effect was more pronounced among patients with current AUD (DiD 0.45 points, 95% CI 0.30 to 0.59; p<0.0001), and those reporting hazardous drinking at baseline (DiD 0.34 points, 95% CI 0.17 to 0.51; p<0.0001; Figure 3).

## Discussion

We present a novel method that integrates genomic and EHR data to identify and evaluate potential drug repurposing candidates for reducing alcohol consumption, including an exemplar pharmacoepidemologic analysis of baclofen. Our approach follows a structured strategy for selecting and assessing drugs for repurposing to treat AUD. Genetic targets were first identified by mapping genome-wide association study (GWAS) loci to genes and expanding alcohol-related gene networks. FDA-approved drugs that interact with these genes were identified through Open Targets, which leverages curated data from ChEMBL. To refine the candidate pool, drugs were excluded if they were associated with severe adverse effects, intended for short-term or as-needed use, approved to treat severe impairing diseases (e.g., Alzheimer’s), or did not meet the threshold for statistical power. Through this filtering process, the initial set of 195 testable drugs was narrowed to 26 promising candidates for further investigation. Finally, our exemplar analysis of one of the candidates, baclofen, served as a test case and provided evidence of its potential to reduce alcohol consumption.

Identified drugs target proteins encoded by several key AUD-related genes, further demonstrating their potential as therapeutic options. Most notably, medications targeting proteins encoded by *HTR1A*, *HTR1B*, *CACNA1C*, and *ADRA2A* genes were identified. *HTR1A* and *HTR1B* encode the presynaptic inhibitory autoreceptors serotonin 1A and 1B, which are linked to the modulation of alcohol consumption^54,55^. Evidence suggests that activation of these receptors reduces alcohol consumption in animal models, with 5HT1A agonism also showing reductions in humans^55^. The *CACNA1C* gene, which encodes the alpha-1C subunit of L-type voltage-gated calcium channels, has been associated with reduced alcohol-seeking behaviors in preclinical studies following inhibition or gene knockout in rats^56^, an effect mediated by the central amygdala^57^. The *ADRA2A* gene, which encodes the alpha-2A adrenergic receptor, is another promising target, as its antagonism reduces alcohol consumption in mice ^58^. The evidence for the latter target is in line with growing broader evidence on the role of the noradrenergic system toward the development of novel medications for AUD (for review, see^59^).

This approach successfully identified several clinically relevant drugs for evaluation as potential AUD treatments. Specifically, *PDE4B*, which encodes an enzyme involved in cAMP hydrolysis in inflammatory cells, was highlighted. Preclinical research shows that inhibition of this enzyme reduces binge drinking, alcohol intake, and other motivational measures associated with alcohol consumption in animal models^60,61^, though more research is needed to determine efficacy in humans. Some clinical evidence supports the use of apremilast, a PDE4 inhibitor, to reduce alcohol intake^61^. Similarly, ibudilast, a multi-phosphodiesterase and macrophage migration inhibitory factor inhibitor, diminished heavy alcohol-drinking and alcohol cue-included neural activity^62^. Buspirone, a 5HT1A agonist, has similarly demonstrated reductions in alcohol consumption, although much of this research is dated^63,64^. Unexpectedly, DPP-4 inhibitors, a class of drugs approved for diabetes mellitus type 2, emerged as a potential target. These drugs act by blocking GLP-1 degradation and therefore boosting endogenous GLP-1 levels. However, research generally favors GLP-1RAs over DPP-4 inhibitors for alcohol use reduction. GLP-1 treatments have shown significantly greater efficacy, including in a recent side-by-side pharmacoepidemiology analysis using AUDIT-C as the outcome^29^. For a recent review on the potential role of the GLP-1 system toward medication development for AUD, see^65^. This may be a result of the network-based approach that identified a putatively relevant pathway for AUD, but that missed an optimal therapeutic target. Indeed, although targets with genetic and network support are enriched for therapeutic efficacy, efficacy is not demonstrated for most of these and this approach does not identify all possible targets^18–21^. However, FDA-approved AUD treatments such as naltrexone, disulfiram, and acamprosate were also identified, substantiating the validity of our approach.

Our exemplar pharmacoepidemiologic analysis evaluated baclofen for its association with reduced alcohol consumption in the VA EHR. Using robust, generalizable methods, we observed that patients initiating baclofen had a significantly greater decrease in AUDIT-C scores than propensity-score matched unexposed comparators. Although the overall effect size was modest, this is consistent with prior EHR-based studies of AUD treatments—topiramate, for example, showed a small effect despite strong RCT support^26^. Importantly, the largest effect of baclofen was among patients with current AUD and those reporting hazardous consumption levels at baseline; this observation is consistent with the overall notion in the field that people with more severe forms of AUD are more likely to respond to baclofen^66^. A recent meta-analysis of 17 trials indicated that baclofen reduces risk of relapse and increases percentage of abstinent days but does not reduce heavy drinking days or drinks per drinking day^45^. Subgroup analysis in this meta-analysis indicated that baclofen is effective in patients who have been withdrawn from alcohol but not in those who are still drinking at the start of treatment. In contrast, we found baclofen is associated with reduced alcohol use in patients who are currently drinking. Although more mechanistic research is needed, prior studies suggest that baclofen may work by amplifying subjective effects of alcohol^67^ and/or dissociating the link between an initial drink and further alcohol^68^ in currently drinking patients. Based on our findings and prior research, larger scale clinical trials are needed to investigate baclofen’s efficacy in moderating drinking comparing it to FDA-approved drugs (e.g., naltrexone).

Our approach to identifying candidates for repurposing in the treatment of AUD has limitations. First, while this method identifies promising repurposing candidates, its reliance on VA EHR data, which is not publicly accessible, may constrain broader implementation.

However, similar approaches could be applied in other large, integrated health systems with rich EHR data. Second, the pharmacoepidemiologic component is best suited to evaluate repurposing candidates that are already in clinical use and for which sufficient exposure and alcohol consumption data exist rather than testing completely novel agents. Third, although these analyses estimate robust associations rather than definitive causal effects, they can still help prioritize candidates for subsequent mechanistic work and randomized trials. Fourth, we only selected one of 20 alcohol-related networks to prioritize a manageable number of promising targets. As a result, it is probable that disease relevant proteins exist within the other 19 networks. Alternative innovative target prioritization and network-based methods should be considered in future work^69^. Furthermore, updating network-based analyses with the largest PAU and related GWAS (e.g., alcohol consumption, general addiction risk) will be essential^70,71^.

We have presented a novel method for identifying drug repurposing opportunities by integrating genomic insights with large-scale real-world evidence. This approach facilitates the prioritization of potential therapeutic candidates for AUD, a condition that continues to struggle from suboptimal treatment options. The potential benefit of our methodology may become even more pronounced as more genetic variants associated with alcohol use are identified, and as our pharmacoepidemiologic findings converge with clinical trial evidence and are replicated across other healthcare systems. Furthermore, it may serve as a generalizable framework for repurposing efforts across other neuropsychiatric conditions. Future work building upon our methodology, including utilizing alternative gene network algorithms or drug filtering processes, is needed to further understand the full potential benefit of our novel method.

## Data Availability

This study is a secondary analysis of existing datasets. Individual-level electronic health record data are not publicly available due to privacy restrictions. Publicly available omics data used in this study can be accessed from the repositories cited in the manuscript.

## Disclaimer

The contents of this publication are the sole responsibility of the authors and do not necessarily reflect the official policy or position of the Uniformed Services University of the Health Sciences, the Department of Defense, or Henry M. Jackson Foundation for the Advancement of Military Medicine.

## Conflict of Interest Disclosures

Dr. Fiellin’s wife is Founder of Playbl, Inc., which disseminates serious videogames for substance use prevention. Dr. Leggio reported receiving honoraria from the UK Medical Council on Alcohol for his work as Editor-in-Chief for the journal Alcohol and Alcoholism and receiving royalties from Routledge for a published textbook. Dr. Kranzler is a member of advisory boards for Altimmune and Clearmind Medicine; a consultant to Sobrera Pharmaceuticals and Altimmune; the recipient of research funding and medication supplies for an investigator-initiated study from Alkermes; a member of the American Society of Clinical Psychopharmacology’s Alcohol Clinical Trials Initiative, which was supported in the last three years by Alkermes, Dicerna, Ethypharm, Imbrium, Indivior, Kinnov, Lilly, Otsuka, and Pear; and an inventor on U.S. provisional patent “Multi-ancestry Genome-wide Association Meta-analysis of Buprenorphine Treatment Response.” All other authors report no biomedical financial interests or potential conflicts of interest.

## Funding

Drs. Rentsch, Kranzler, and Gray are supported by NIAAA grant R01 AA030041. Drs. Farokhnia and Leggio are supported by the NIH Intramural Research Program (NIDA and NIAAA). Drs. Justice and Fiellin are supported by NIAAA grant and P01 AA029545. Dr. Fiellin is supported by NIAAA grant R01 AA023733. Dr. Kranzler is supported by the Veterans Integrated Service Network’s Mental Illness Research, Education and Clinical Center; U.S. Department of Veterans Affairs grant I01 BX004820 and NIAAA grant R01 AA030056. Dr. Gray is supported by Department of Defense grant HU0001-22-2-0066.

## Author contributions

Study concept and design: JCG, CTR, HRK. Analysis of data: CTR, ZP, JCG, ELM. Drafting of the paper: SGM, CTR, JCG, HRK. Critical revision of the paper for important intellectual content: MS, MRS, MF, JT, ACJ, DAF, LL.

## Notes

### Competing Interest Statement

The wife of Dr. Fiellin is the Founder of Playbl, Inc., which disseminates serious videogames for substance use prevention. Dr. Leggio reported receiving honoraria from the UK Medical Council on Alcohol for his work as Editor-in-Chief for the journal Alcohol and Alcoholism and receiving royalties from Routledge for a published textbook. Dr. Kranzler is a member of advisory boards for Altimmune and Clearmind Medicine; a consultant to Sobrera Pharmaceuticals and Altimmune; the recipient of research funding and medication supplies for an investigator-initiated study from Alkermes; a member of the American Society of Clinical Psychopharmacology Alcohol Clinical Trials Initiative, which was supported in the last three years by Alkermes, Dicerna, Ethypharm, Imbrium, Indivior, Kinnov, Lilly, Otsuka, and Pear; and an inventor on U.S. provisional patent: Multi-ancestry Genome-wide Association Meta-analysis of Buprenorphine Treatment Response.  All other authors report no biomedical financial interests or potential conflicts of interest.

### Author Declarations

The Department of Veterans Affairs (VA) study was approved by the institutional review boards of Yale University (ref #1506016006) and VA Connecticut Healthcare System (ref #AJ0013) with a waiver of informed consent.

## References

1. American Psychiatric Association. Diagnostic and Statistical Manual of Mental Disorders . (American Psychiatric Association, 2013).

2. Substance Abuse and Mental Health Services Administration. Center for Behavioral Health Statistics and Quality. 2022 National Survey on Drug Use and Health. Table 5.9B—Alcohol use disorder in past year: among people aged 12 or older; by age group and demographic characteristics, percentages, 2022 and 2023. Center for Behavioral Health Statistics and Quality, Substance Abuse and Mental Health Services Administration (2024).

3. Substance Abuse and Mental Health Services Administration. Center for Behavioral Health Statistics and Quality. 2022 National Survey on Drug Use and Health. Table 5.9A—Alcohol use disorder in past year: among people aged 12 or older; by age group and demographic characteristics, numbers in thousands, 2022 and 2023. Center for Behavioral Health Statistics and Quality, Substance Abuse and Mental Health Services Administration (2024).

4. Han, B., Jones, C. M., Einstein, E. B., Powell, P. A. & Compton, W. M. Use of Medications for Alcohol Use Disorder in the US: Results from the 2019 National Survey on Drug Use and Health. JAMA Psychiatry 78, (2021).

5. Anton, R. F., Schacht, J. P. & Book, S. W. Pharmacologic treatment of alcoholism. in Handbook of Clinical Neurology vol. 125 (2014).

6. Fairbanks, J. et al. Evidence-Based Pharmacotherapies for Alcohol Use Disorder: Clinical Pearls. Mayo Clinic Proceedings vol. 95 Preprint at 10.1016/j.mayocp.2020.01.030 (2020).

7. Müller, C. A., Geisel, O., Banas, R. & Heinz, A. Current pharmacological treatment approaches for alcohol Dependence. Expert Opin Pharmacother 15, (2014).

8. Palpacuer, C. et al. Pharmacologically controlled drinking in the treatment of alcohol dependence or alcohol use disorders: a systematic review with direct and network meta-analyses on nalmefene, naltrexone, acamprosate, baclofen and topiramate. Addiction 113, (2018).

9. Ray, L. A., Green, R. J., Roche, D. J. O., Magill, M. & Bujarski, S. Naltrexone effects on subjective responses to alcohol in the human laboratory: A systematic review and meta-analysis. Addiction Biology 24, (2019).

10. Mark, T. L. et al. Barriers to the use of medications to treat alcoholism. American Journal on Addictions 12, (2003).

11. Ehrie, J., Hartwell, E. E., Morris, P. E., Mark, T. L. & Kranzler, H. R. Survey of Addiction Specialists’ Use of Medications to Treat Alcohol Use Disorder. Front Psychiatry 11, (2020).

12. Wouters, O. J., McKee, M. & Luyten, J. Estimated Research and Development Investment Needed to Bring a New Medicine to Market, 2009-2018. JAMA - Journal of the American Medical Association 323, (2020).

13. DiMasi, J. A., Grabowski, H. G. & Hansen, R. W. Innovation in the pharmaceutical industry: New estimates of R&D costs. J Health Econ 47, (2016).

14. Congressional Budget Office. Research and development in the pharmaceutical industry. Congressional Budget Office Nonpartisan Analysis for the U.S. Congress (2021).

15. Zong, N., et al. Computational drug repurposing based on electronic health records: a scoping review. NPJ Digit Med 5, (2022).

16. Kulkarni, V. S., Alagarsamy, V., Solomon, V. R., Jose, P. A. & Murugesan, S. Drug Repurposing: An Effective Tool in Modern Drug Discovery. Russ J Bioorg Chem 49, (2023).

17. Wang, L. et al. The landscape of the methodology in drug repurposing using human genomic data: a systematic review. Brief Bioinform 25, (2024).

18. King, E. A., Wade Davis, J. & Degner, J. F. Are drug targets with genetic support twice as likely to be approved? Revised estimates of the impact of genetic support for drug mechanisms on the probability of drug approval. PLoS Genet 15, (2019).

19. Minikel, E. V., Painter, J. L., Dong, C. C. & Nelson, M. R. Refining the impact of genetic evidence on clinical success. Nature 629, 624–629 (2024).

20. Barrio-Hernandez, I. et al. Network expansion of genetic associations defines a pleiotropy map of human cell biology. Nat Genet 55, (2023).

21. MacNamara, A. et al. Network and pathway expansion of genetic disease associations identifies successful drug targets. Sci Rep 10, (2020).

22. Austin, P. C. An introduction to propensity score methods for reducing the effects of confounding in observational studies. Multivariate Behav Res 46, (2011).

23. Wang, S. V. et al. Emulation of Randomized Clinical Trials with Nonrandomized Database Analyses: Results of 32 Clinical Trials. JAMA 329, (2023).

24. Farokhnia, M. et al. Spironolactone as a potential new pharmacotherapy for alcohol use disorder: convergent evidence from rodent and human studies. Mol Psychiatry 27, (2022).

25. Rentsch, C. T., Fiellin, D. A., Bryant, K. J., Justice, A. C. & Tate, J. P. Association Between Gabapentin Receipt for Any Indication and Alcohol Use Disorders Identification Test—Consumption Scores Among Clinical Subpopulations With and Without Alcohol Use Disorder. Alcohol Clin Exp Res 43, (2019).

26. Kranzler, H. R. et al. Association of topiramate prescribed for any indication with reduced alcohol consumption in electronic health record data. Addiction 117, (2022).

27. Fluyau, D., Kailasam, V. K. & Pierre, C. G. A Bayesian meta-analysis of topiramate’s effectiveness for individuals with alcohol use disorder. Journal of Psychopharmacology 37, (2023).

28. Kranzler, H. R., Feinn, R., Morris, P. & Hartwell, E. E. A meta-analysis of the efficacy of gabapentin for treating alcohol use disorder. Addiction 114, (2019).

29. Farokhnia, M. et al. Glucagon-like peptide-1 receptor agonists but not dipeptidyl peptidase-4 inhibitors reduce alcohol intake. J Clin Invest 10.1172/JCI188314 (2025) doi: 10.1172/JCI188314.

30. U.S. Department of Veterans Affairs. About the Department. U.S. Department of Veterans Affairs.

31. Zhou, H. et al. Multi-ancestry study of the genetics of problematic alcohol use in over 1 million individuals. Nat Med 29, (2023).

32. Ochoa, D. et al. The next-generation Open Targets Platform: reimagined, redesigned, rebuilt. Nucleic Acids Res 51, (2023).

33. Ji, C., Kerrebijn, I., Arbabi, K., Schipper, M. & Wainberg, M. Benchmarking genome-wide association study causal gene prioritization for drug discovery. medRxiv 2025.09.23.25336370 (2025) doi:10.1101/2025.09.23.25336370.

34. Mountjoy, E. et al. An open approach to systematically prioritize causal variants and genes at all published human GWAS trait-associated loci. Nat Genet 53, (2021).

35. Mendez, D. et al. ChEMBL: Towards direct deposition of bioassay data. Nucleic Acids Res 47, (2019).

36. Papadatos, G., Gaulton, A., Hersey, A. & Overington, J. P. Activity, assay and target data curation and quality in the ChEMBL database. J Comput Aided Mol Des 29, (2015).

37. Zdrazil, B. et al. The ChEMBL Database in 2023:ã drug disco v ery platf orm spanning multiple bioactivity data typesãnd time periods. Nucleic Acids Res 52, (2024).

38. Fang, H. et al. FDALabel for drug repurposing studies and beyond. Nature Biotechnology vol. 38 Preprint at 10.1038/s41587-020-00751-0 (2020).

39. Kuhn, M., Letunic, I., Jensen, L. J. & Bork, P. The SIDER database of drugs and side effects. Nucleic Acids Res 44, (2016).

40. Bush, K., Kivlahan, D. R., McDonell, M. B., Fihn, S. D. & Bradley, K. A. The AUDIT alcohol consumption questions (AUDIT-C): an effective brief screening test for problem drinking. Ambulatory Care Quality Improvement Project (ACQUIP). Alcohol Use Disorders Identification Test. Arch Intern Med 158, (1998).

41. Fiellin, D. A., Reid, M. C. & O’Connor, P. G. Screening for alcohol problems in primary care: A systematic review. Arch Intern Med 160, (2000).

42. Kranzler, H. R. & Hartwell, E. E. Medications for treating alcohol use disorder: A narrative review. Alcohol: Clinical and Experimental Research vol. 47 Preprint at 10.1111/acer.15118 (2023).

43. Agabio, R. et al. Baclofen for the treatment of alcohol use disorder: the Cagliari Statement. Lancet Psychiatry 5, 957–960 (2018).

44. Rolland, B., Simon, N., Franchitto, N. & Aubin, H. J. France grants an approval to baclofen for alcohol dependence. Alcohol and Alcoholism vol. 55 Preprint at 10.1093/alcalc/agz082 (2020).

45. Agabio, R., Saulle, R., Rösner, S. & Minozzi, S. Baclofen for alcohol use disorder. Cochrane Database of Systematic Reviews 1, (2023).

46. Bush, K., Kivlahan, D. R., McDonell, M. B., Fihn, S. D. & Bradley, K. A. The AUDIT alcohol consumption questions (AUDIT-C): An effective brief screening test for problem drinking. Arch Intern Med 158, (1998).

47. Brookhart, M. A. et al. Variable selection for propensity score models. Am J Epidemiol 163, (2006).

48. Pearl, J. Invited commentary: Understanding bias amplification. American Journal of Epidemiology vol. 174 Preprint at 10.1093/aje/kwr352 (2011).

49. Cormen, T. H., Leiserson, C. E. & Rivest, R. L. Introduction to Algorithms , Second Edition. Computer vol. 7 (2001).

50. Donald, S. G. & Lang, K. Inference with difference-in-differences and other panel data. Review of Economics and Statistics 89, (2007).

51. Lechner, M. The Estimation of Causal Effects by Difference-in-Difference Methods. Foundations and Trends in Econometrics vol. 4 (2010).

52. Witkiewitz, K., Litten, R. Z. & Leggio, L. Advances in the science and treatment of alcohol use disorder. Sci Adv 5, (2019).

53. Perry, C. et al. The Management of Substance Use Disorders: Synopsis of the 2021 U.S. Department of Veterans Affairs and U.S. Department of Defense Clinical Practice Guideline. Ann Intern Med 175, (2022).

54. Belmer, A., Depoortere, R., Beecher, K., Newman-Tancredi, A. & Bartlett, S. E. Neural serotonergic circuits for controlling long-term voluntary alcohol consumption in mice. Mol Psychiatry 27, (2022).

55. Castle, M. E. & Flanigan, M. E. The role of brain serotonin signaling in excessive alcohol consumption and withdrawal: A call for more research in females. Neurobiol Stress 30, (2024).

56. Uhrig, S. et al. Differential Roles for L-Type Calcium Channel Subtypes in Alcohol Dependence. Neuropsychopharmacology 42, (2017).

57. Varodayan, F. P., De Guglielmo, G., Logrip, M. L., George, O. & Roberto, M. Alcohol dependence disrupts amygdalar L-type voltage-gated calcium channel mechanisms. Journal of Neuroscience 37, (2017).

58. Micioni Di Bonaventura, M. V., et al. Effects of A 2A adenosine receptor blockade or stimulation on alcohol intake in alcohol-preferring rats. Psychopharmacology (Berl) 219, (2012).

59. Haass-Koffler, C. L., Swift, R. M. & Leggio, L. Noradrenergic targets for the treatment of alcohol use disorder. Psychopharmacology (Berl) 235, (2018).

60. Leonardo Jimenez Chavez, C., Bryant, C. D., Munn-chernoff, M. A. & Szumlinski, K. K. Selective inhibition of pde4b reduces binge drinking in two c57bl/6 substrains. Int J Mol Sci 22, (2021).

61. Grigsby, K. B. et al. Preclinical and clinical evidence for suppression of alcohol intake by apremilast. Journal of Clinical Investigation 133, (2023).

62. Grodin, E. N. et al. Ibudilast, a neuroimmune modulator, reduces heavy drinking and alcohol cue-elicited neural activation: a randomized trial. Transl Psychiatry 11, (2021).

63. Kranzler, H. R. et al. Buspirone Treatment of Anxious Alcoholics: A Placebo-Controlled Trial. Arch Gen Psychiatry 51, (1994).

64. Malec, T. S., Malec, E. A. & Dongier, M. Efficacy of buspirone in alcohol dependence: A review. Alcohol Clin Exp Res 20, (1996).

65. Bruns VI, N., Tressler, E. H., Vendruscolo, L. F., Leggio, L. & Farokhnia, M. IUPHAR review – Glucagon-like peptide-1 (GLP-1) and substance use disorders: An emerging pharmacotherapeutic target. Pharmacol Res 207, 107312 (2024).

66. Leggio, L., Garbutt, J. & Addolorato, G. Effectiveness and Safety of Baclofen in the Treatment of Alcohol Dependent Patients. CNS Neurol Disord Drug Targets 9, (2012).

67. Farokhnia, M. et al. Biobehavioral effects of baclofen in anxious alcohol-dependent individuals: A randomized, double-blind, placebo-controlled, laboratory study. Transl Psychiatry 7, (2017).

68. Farokhnia, M. et al. A deeper insight into how GABA-B receptor agonism via baclofen may affect alcohol seeking and consumption: lessons learned from a human laboratory investigation. Mol Psychiatry 26, (2021).

69. Chen, R., Duffy, Á. & Do, R. Genomics of drug target prioritization for complex diseases. Nat Rev Genet 10.1038/S41576-025-00904-4 (2025) doi: 10.1038/S41576-025-00904-4.

70. Saunders, G. R. B. et al. Genetic diversity fuels gene discovery for tobacco and alcohol use. Nature 612, 720–724 (2022).

71. Hatoum, A. S. et al. Multivariate genome-wide association meta-analysis of over 1 million subjects identifies loci underlying multiple substance use disorders. Nature. Mental health 1, 210–223 (2023).

