## Supplementary material for "Bridging Genomics and Pharmacoepidemiology to Expand Treatment Options for Alcohol Use Disorder": Tables and Figures

**Table 1.** Drug repurposing candidates for alcohol use disorder

| **Drug*** | **Gene(s)** | **Source** | **Indication(s)** | **VA patients with ≥1 prescription and AUDIT-C > 0** |
| --- | --- | --- | --- | --- |
| Pregabalin | *CACNA1C*, *CACNA1D*, *CACNB2*, *CACNB3* | Network/PAU | Neuropathic pain, neuralgia, epilepsy, fibromyalgia | 102,043 |
| Gabapentin | *CACNA1C*, *CACNA1D*, *CACNB2*, *CACNB3* | Network/PAU | Neuralgia, epilepsy, restless legs syndrome | 927,239 |
| Clonidine | *ADRA2A*, *ADRA2C*, *ADRA2B* | Network | Attention-deficit/hyperactivity disorder, hypertension, neuropathic pain, glaucoma, migraine | 89,720 |
| Guanfacine | *ADRA2A*, *ADRA2C*, *ADRA2B* | Network | Attention-deficit/hyperactivity disorder, hypertension | 2,669 |
| Labetalol | *ADRA2A*, *ADRA2C*, *ADRA2B* | Network | Hypertension, cardiovascular disease | 10,619 |
| Mirtazapine | *ADRA2A*, *ADRA2C*, *ADRA2B* | Network | Depression, insomnia | 328,222 |
| Tizanidine | *ADRA2A*, *ADRA2C*, *ADRA2B* | Network | Spasticity | 81,483 |
| Amlodipine | *CACNA1C*, *CACNA1D* | Network/PAU | Hypertension, angina, heart failure | 981,661 |
| Felodipine | *CACNA1C*, *CACNA1D* | Network/PAU | Hypertension, angina | 7,739 |
| Nifedipine | *CACNA1C*, *CACNA1D* | Network/PAU | Hypertension, angina, vasospasm | 79,234 |
| Diltiazem | *CACNA1C*, *CACNA1D* | Network/PAU | Hypertension, angina, arrhythmias | 130,502 |
| Verapamil | *CACNA1C*, *CACNA1D* | Network/PAU | Hypertension, angina, arrhythmias | 42,120 |
| Alogliptin | *DPP4* | Network | Type 2 diabetes mellitus | 111,929 |
| Linagliptin | *DPP4* | Network | Type 2 diabetes mellitus | 3,746 |
| Saxagliptin | *DPP4* | Network | Type 2 diabetes mellitus | 52,077 |
| Sitagliptin | *DPP4* | Network | Type 2 diabetes mellitus, hyperlipidemia, cardiovascular disease | 8,929 |
| Buspirone | *HTR1A* | Network | Anxiety, depression | 231,543 |
| Vilazodone | *HTR1A* | Network | Depression | 4,499 |
| Vortioxetine | *HTR1A* | Network | Depression | 5,008 |
| Zolmitriptan | *HTR1B* | Network | Migraine | 21,302 |
| Eletriptan | *HTR1B*, *HTR1D* | Network | Migraine | 9,781 |
| Naratriptan | *HTR1B* | Network | Migraine | 446 |
| Almotriptan | *HTR1B*, *HTR1D* | Network | Migraine | 251 |
| Baclofen | *GABBR2*, *GABBR1* | Network | Spasticity, multiple sclerosis, cerebral palsy, spinal cord injury | 151,361 |
| Roflumilast | *PDE4B* | PAU | Chronic obstructive pulmonary disease, chronic bronchitis | 4,112 |
| Apremilast | *PDE4B* | PAU | Psoriasis, psoriatic arthritis, oral ulcers | 3,767 |

*Note*. Drugs not separated by a line were consolidated into medication groups for testing based on identical or near-identical mechanisms of action and treatment indications. Genes were identified by the problematic alcohol use (PAU) genome-wide association study and the top alcohol-related network (Network). AUDIT-C = Alcohol Use Disorders Identification Test-Consumption, VA = Veterans Affairs.

**Table 2.** Estimated average pre- and post-index date AUDIT-C scores and difference-in-differences (DiD), overall, by baseline AUD and level of alcohol consumption

|  |  | **Exposed** | **Unexposed** |
| --- | --- | --- | --- |
|  |  | **n=22,295** | **n=18,180** |
| **All patients** | **Pre** | 2.97 (0.02) | 2.98 (0.02) |
|  | **Post** | 2.09 (0.02) | 2.29 (0.02) |
|  | **D^n^** | -0.88 (0.02) | -0.69 (0.03) |
|  | **DiD (95% CI)** | 0.19 (0.12, 0.26), p<0.0001 | |
| **By baseline AUD** | |  |  |
| No AUD |  | **n=17,618** | **n=14,400** |
|  | **Pre** | 2.32 (0.02) | 2.33 (0.02) |
|  | **Post** | 1.70 (0.02) | 1.84 (0.02) |
|  | **D^n^** | -0.62 (0.03) | -0.50 (0.03) |
|  | **DiD (95% CI)** | 0.12 (0.04, 0.20), p=0.0018 | |
| AUD |  | **n=4,677** | **n=3,780** |
|  | **Pre** | 5.40 (0.04) | 5.43 (0.04) |
|  | **Post** | 3.53 (0.04) | 4.02 (0.04) |
|  | **D^n^** | -1.86 (0.05) | -1.42 (0.06) |
|  | **DiD (95% CI)** | 0.45 (0.30, 0.59), p<0.0001 | |
| **By baseline AUDIT-C** | |  |  |
| Low-risk |  | **n=15,994** | **n=13,158** |
|  | **Pre** | 1.62 (0.01) | 1.65 (0.02) |
|  | **Post** | 1.44 (0.01) | 1.60 (0.02) |
|  | **D^n^** | -0.18 (0.02) | -0.05 (0.02) |
|  | **DiD (95% CI)** | 0.13 (0.07, 0.19), p<0.0001 | |
| At-risk |  | **n=4,392** | **n=3,487** |
|  | **Pre** | 4.82 (0.03) | 4.85 (0.03) |
|  | **Post** | 3.20 (0.03) | 3.52 (0.03) |
|  | **D^n^** | -1.62 (0.04) | -1.33 (0.04) |
|  | **DiD (95% CI)** | 0.29 (0.18, 0.41), p<0.0001 | |
| Hazardous/binge | | **n=1,909** | **n=1,535** |
|  | **Pre** | 9.97 (0.04) | 10.12 (0.05) |
|  | **Post** | 4.93 (0.04) | 5.42 (0.05) |
|  | **D^n^** | -5.04 (0.06) | -4.70 (0.06) |
|  | **DiD (95% CI)** | 0.34 (0.17, 0.51), p=0.0001 | |
| *Note*. Average pre- and post-index date AUDIT-C scores reported as mean (standard error). AUDIT-C = Alcohol Use Disorders Identification Test - Consumption; AUD = alcohol use disorder; Pre - pre-index AUDIT-C score; Post - post-index AUDIT-C score; D^n^ - change in AUDIT-C score; CI - confidence interval | | | |

**Figure 1.** Study pipeline. (1a) Loci from a GWAS of problematic alcohol use (PAU) were assigned to genes based on their presence within the gene or proximity to the nearest transcription start site. Reproduced from Fig. 1f of ^31^. (1b) The top alcohol-related gene network was derived from the output of a recently developed and validated method for network-based expansion of causal genes. Reproduced from Fig. 1c of ^20^. (2) Genes were linked to medications using the Open Targets Platform ^32^, with data curated to include only FDA-approved drugs. (3) FDA-approved drugs linked to these genes were filtered based on severe adverse effects, short-term or as-needed use, approval for treating severe impairing diseases, or not meeting the threshold for statistical power. Drugs with similar mechanisms of action were consolidated into medication groups. (4) Pharmacoepidemiologic analysis of the candidate drug versus propensity-score matched patients in the VA EHR. The exemplar candidate drug reported in this study was baclofen.

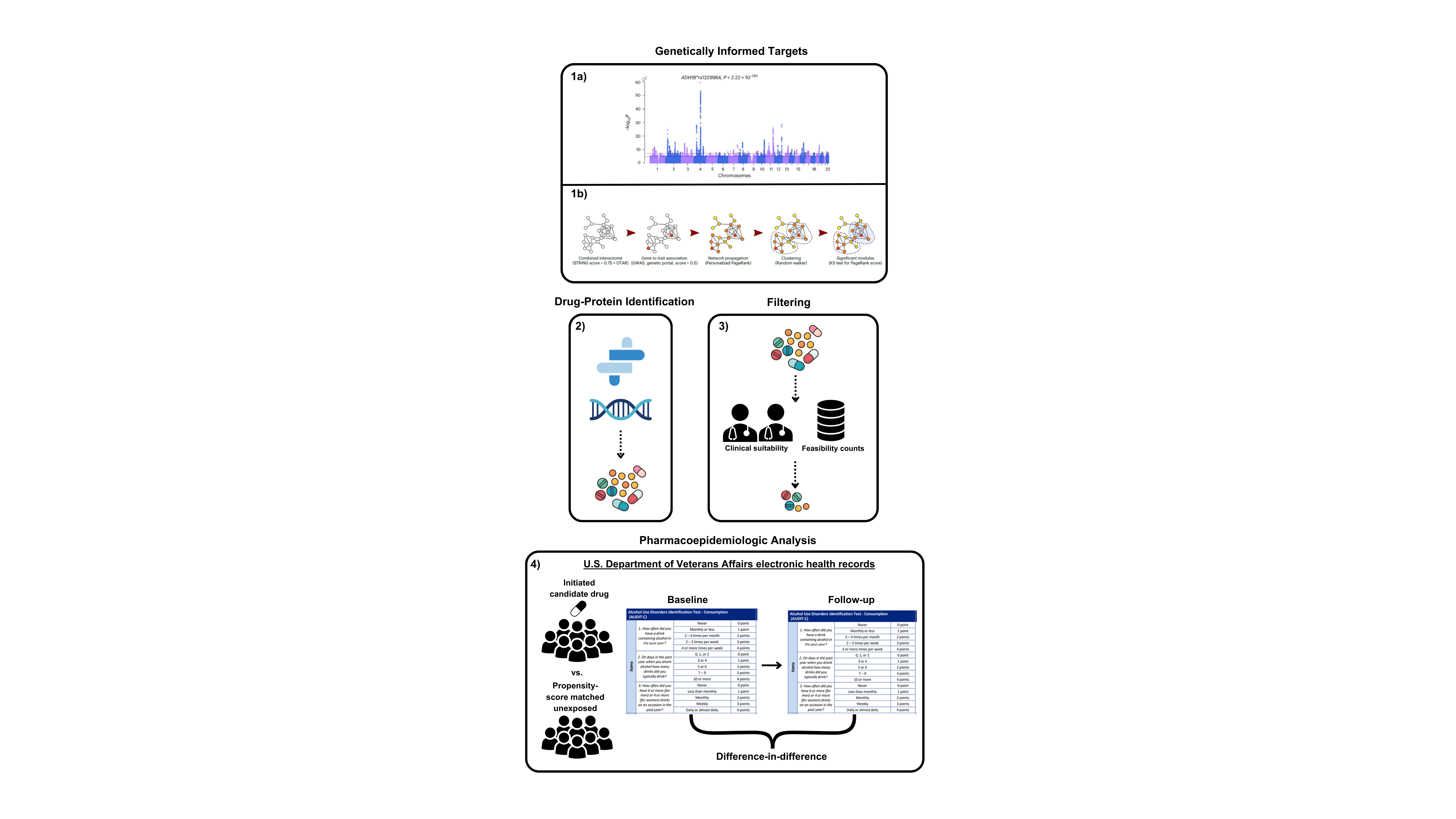

**Figure 2.** Diagram depicting the flow of the pipeline.

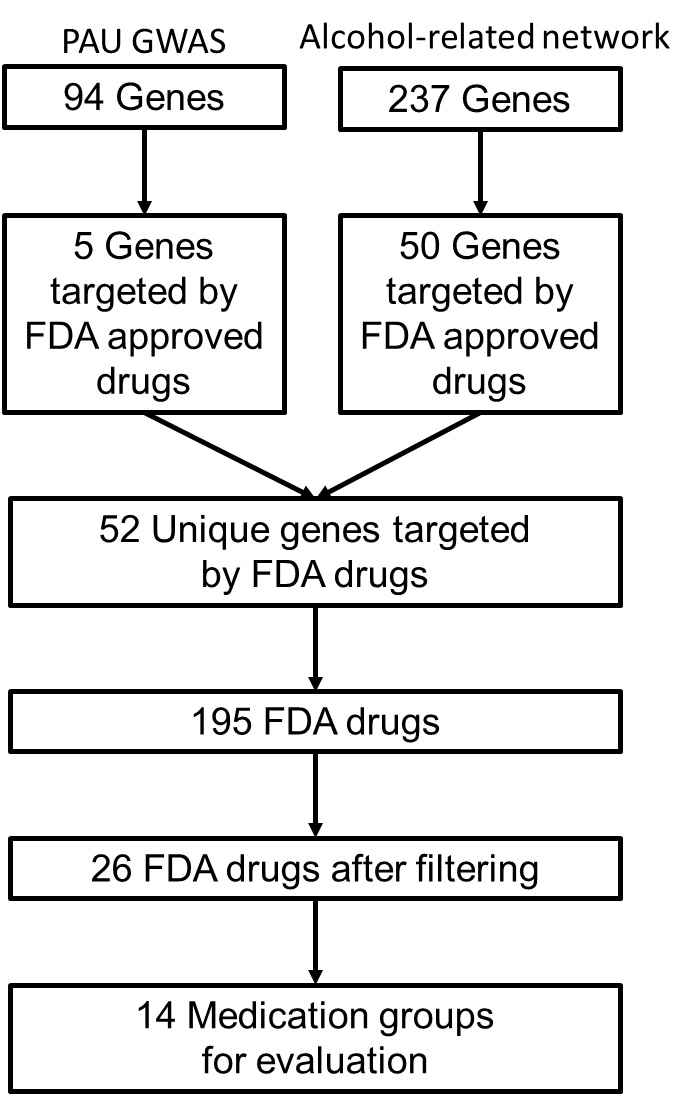

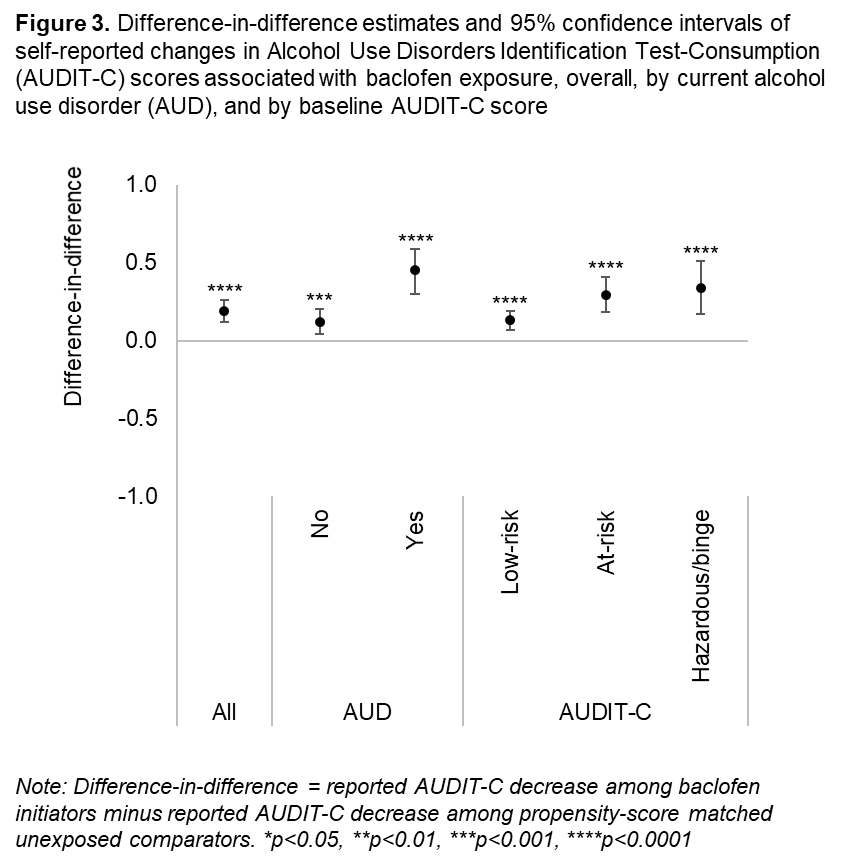
